# An AI-assisted feasibility evaluation of three photoplethysmography-derived microvascular reactivity signals in MIMIC-IV-WDB v0.1.0

**DOI:** 10.64898/2026.06.03.26354863

**Authors:** Thomas C. Landry, Youjin Kim

## Abstract

**Background:** Capillary refill time, an examiner-dependent bedside test of distal microvascular perfusion, has become a resuscitation target in septic shock, ^1,2,3,4^ motivating a continuous surrogate computed from the photoplethysmogram (PPG, the optical waveform the pulse oximeter on every ICU patient already records). ^5,6,7,8^

**Objective:** We attempted three PPG-derived candidate measures on the MIMIC-IV Waveform Database (MIMIC-IV-WDB v0.1.0) and asked, by inspecting randomly drawn examples, whether each captured its intended physiology before any downstream modeling.

**Methods:** MIMIC-IV-WDB v0.1.0^9^ was linked to MIMIC-IV. ^10^ The signals were a cuff-anchored perfusion-index recovery (reactive hyperemia when the cuff shares an arm with the probe), a slow Mayer-wave-band power ratio of the perfusion index (sympathetic vasomotor tone), and a per-beat diastolic exponential decay time constant (a refill-like recovery time). For each signal we drew 10 random examples at a fixed seed and checked them against a checklist fixed in advance. Each was read by the author and, separately, by MedGemma 1.5, a multimodal medical language model run locally. A synthetic test with a known time constant checked the third signal.

**Results:** The cuff-anchored signal showed the expected occlusion-reperfusion shape on 268 of 6,236 evaluable cuff cycles (4.30%) in 15 of 19 patients, consistent with opposite-limb placement of the probe and cuff. The slow-band ratio returned a stable cohort value, but a clear, stationary peak appeared in only 4 of 10 random windows. The per-beat fit met its goodness-of-fit threshold in 10 of 10 beats, yet a cardiac-frequency heuristic flagged a possible fit on the heart-rate oscillation in 7 of 10, and in 5 of 17 patients the time constant lay where an exponential is indistinguishable from a straight line. A 0.5 Hz high-pass pre-filter implanted its own approximately 318 ms time constant regardless of truth. The language model tracked the human on clear positives but reported the pattern present on every call it returned, never absent.

**Conclusions:** Two of the three candidate signals did not reflect their intended physiology in most examples, and the third was constrained by sensor placement. Inspecting a few random raw inputs against a checklist written in advance is an inexpensive upstream check before downstream inference on PPG-derived microvascular signals.

## 1. Introduction

### 1.1. Capillary refill time as a resuscitation target

Capillary refill time (CRT) is a bedside test of distal microvascular perfusion. The examiner presses on a fingertip, releases, and counts the seconds it takes for normal color to return. A prolonged refill is a sign that small blood vessels are not delivering flow normally, and in critically ill adults it carries prognostic weight. ANDROMEDA-SHOCK compared lactate-guided with refill-guided resuscitation in adults with septic shock and reported a nonsignificant reduction in 28-day mortality with a clearer reduction in organ dysfunction in the refill arm. ^1^ A Bayesian reanalysis of those data estimated a high posterior probability of a real mortality benefit at clinically reasonable priors. ^2^ ANDROMEDA-SHOCK-2 then randomized 1,467 patients with early septic shock to a personalized hemodynamic protocol targeting capillary refill versus usual care, reporting superiority on a hierarchical composite of mortality, vital support duration, and length of stay (win ratio 1.16, 95% confidence interval 1.02 to 1.33). ^3^ A 2023 systematic review with meta-analysis confirms that prolonged refill predicts mortality across heterogeneous critical-illness cohorts. ^4^

### 1.2. The measurement-quality problem

The clinical case for capillary refill is therefore stronger than it has ever been, but the test itself remains examiner-dependent. A European intensivist survey found wide between-examiner variability in compression force, duration, anatomic site, and what counts as a positive result. ^11^ A 2024 methods review enumerates more than ten device-assisted alternatives that have been proposed, none of which has reached routine bedside use. ^12^ A continuous, examiner-independent capillary-refill analog computed from a signal that ICU monitors already record would address this gap, and the pulse oximeter is the most accessible substrate.

### 1.3. The photoplethysmogram and the perfusion index

The pulse oximeter shines red and infrared light through a fingertip and measures how much is absorbed by the tissue and blood. The resulting time-varying light absorption is called the photoplethysmogram (PPG). It rises and falls with each heartbeat as more or less arterial blood passes under the sensor. From the PPG, monitors derive the saturation reading (SpO_2_) and a less widely used value called the perfusion index (PI), which is the ratio of the pulsatile component of the signal to the non-pulsatile component, expressed as a percent. The perfusion index is therefore a continuous, monitor-side measure of how much flow is reaching the fingertip; a low or falling PI is consistent with peripheral hypoperfusion and a high or rising PI with peripheral vasodilation. ^23^

### 1.4. Prior PPG-derived microvascular signals

Several groups have proposed PPG-derived signals as continuous surrogates for capillary refill or microvascular reactivity. Hunter and colleagues trained a supervised classifier on PPG morphology to detect flash and prolonged refill against bedside reference. ^5^ Yamamoto and colleagues defined a quantitative pulse-oximetry refill measure (Q-CRT) in liver-transplant recipients. ^6^ Sundrani and colleagues used PPG and electrocardiogram features to predict emergency-to-ICU decompensation in a large multi-site cohort. ^7^ Yin and colleagues used PPG morphology features overlapping with refill decay shape to estimate noninvasive blood pressure. ^8^ These works generally evaluate a proposed signal against a downstream label (manual CRT, mortality, blood pressure) rather than first confirming that the proposed signal is computing what it is claimed to compute on the underlying waveform. That second, upstream question is the focus of the present work.

### 1.5. The three candidate signals attempted here

We attempted three candidate microvascular-function signals on MIMIC-IV-WDB v0.1.0 (Figure 1). Each starts from the PPG signal that the pulse oximeter already records on every ICU patient.

**Figure 1:**
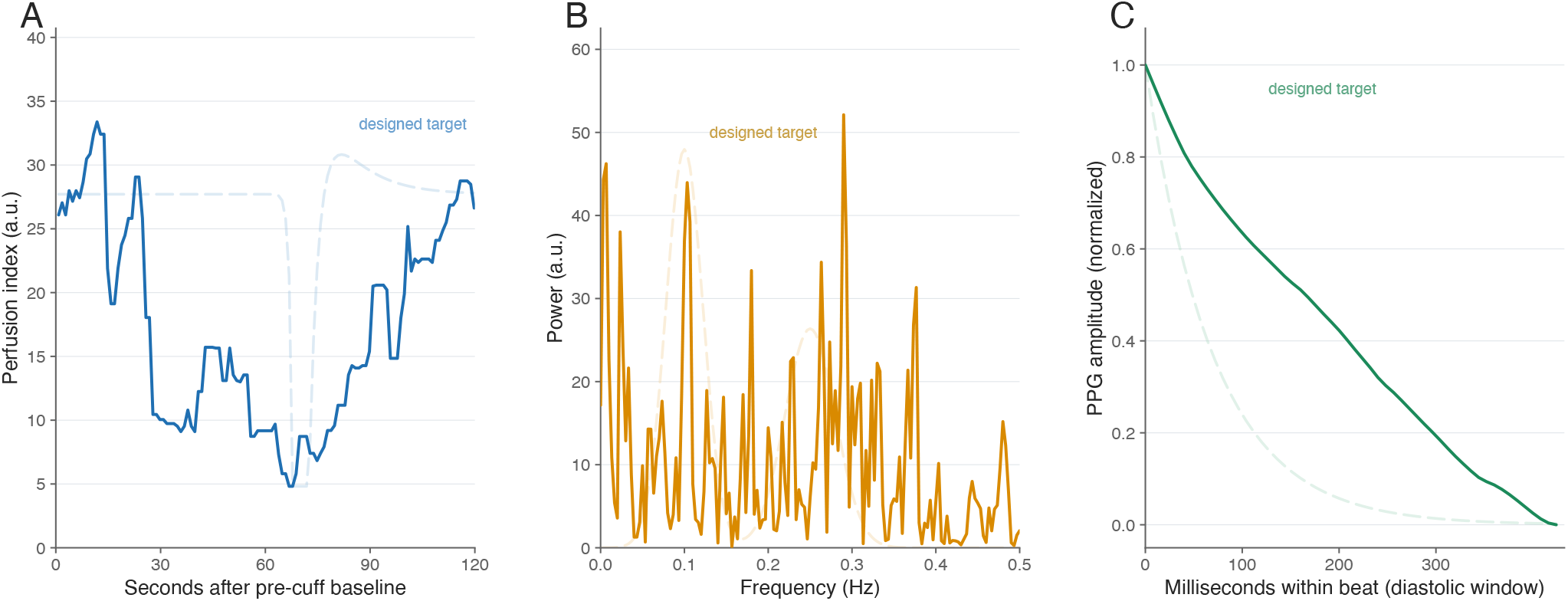
What each candidate signal was designed to capture, and what the data actually show. In every panel the faint dashed curve is the idealized target the method assumes (a model, not measured data) and the solid curve is one real, representative recording on the same axes; the gap between them is the result. **(A)** Signal 1, the cuff-anchored perfusion-index recovery: the textbook reactive-hyperemia target dips at cuff release and overshoots baseline before settling, while the real trace dips and returns without the overshoot. **(B)** Signal 2, the Mayer-band spectrum: the target is a clean low-frequency (Mayer) peak beside a respiratory peak, while the real 5-minute periodogram is broadband and spiky with no dominant peak. **(C)** Signal 3, the per-beat diastolic decay: the target is a clean single exponential, while a real long-time-constant beat is nearly a straight line. Each real trace is the clearest available example for that signal, drawn at a fixed seed; selection criteria are in Methods.

#### Signal 1: cuff-anchored perfusion-index recovery

ICU monitors periodically inflate a blood pressure cuff above systolic pressure, hold, and deflate over 30 to 60 seconds. If the cuff and the pulse oximeter probe happen to be on the same arm, the cuff cycle is, mechanically, a brief arterial occlusion of the same arm whose recovery on cuff release the PPG records. Distal to the cuff, vasodilators accumulate during the brief ischemia and the dilated vascular bed accommodates a transient flow overshoot called reactive hyperemia. ^13,14,15,16,17,18,24^ If the perfusion-index trace after cuff deflation shows the expected occlusion-and-recovery pattern, the recovery slope is a candidate microvascular-reactivity signal.

#### Signal 2: Mayer-band power ratio of the perfusion-index envelope

Mayer waves are slow, spontaneous oscillations near 0.1 Hz (one cycle every 10 seconds) that have been observed in arterial pressure, muscle sympathetic nerve activity, peripheral vascular resistance, and skin blood flow since the late 19th century. Their amplitude rises and falls with manipulations that change sympathetic drive (orthostatic stress, mental stress, vasoactive drugs), which is the basis for using their power as a noninvasive surrogate for sympathetic vasomotor tone. ^19^ If sympathetic drive imprints these slow oscillations on the perfusion-index envelope, then summarizing how much of the signal’s power sits in the Mayer band is a candidate vasomotor-tone readout. We followed the long-standing heart-rate-variability convention ^20^ of splitting the spectrum into a low-frequency band (LF, 0.05–0.15 Hz, where Mayer waves live) and a high-frequency band (HF, 0.15–0.40 Hz, where respiratory modulation lives) and reporting the normalized ratio *P*_LF_ /( *P*_LF_ + *P*_HF_ ). The ratio is preferred over absolute LF power because it controls for the overall amplitude of the underlying signal and is the conventional summary in this literature.

#### Signal 3: per-beat exponential time constant on the diastolic limb of the PPG

On each heartbeat the PPG rises to a peak in systole and falls back during diastole through a small downstroke called the dicrotic notch (which marks aortic valve closure) before reaching the diastolic minimum just before the next beat. The simplest analytic model of how arterial pressure should fall during diastole is the two-element Windkessel, first proposed by Otto Frank in 1899 as an analogy to the air chamber (German: *Windkessel*) in a fire-engine pump: the major arteries act as an elastic chamber that absorbs flow during systole and discharges it through a peripheral resistance during diastole. Under closed-valve diastole the model predicts pressure decays exponentially as *y* (*t*) = A exp (−*t* / *τ*) + *C*, where the time constant *τ* is the product of total peripheral resistance *R* and lumped arterial compliance *C* and represents the characteristic time over which the pressure falls by a factor of *e* ≈ 2.72. ^23^ We chose a single-exponential model rather than a more elaborate bi-exponential or higher-order Windkessel because the diastolic limb of a fingertip PPG beat is short (typically 300–600 ms in adults), and a one-parameter passive-decay summary is the simplest model that the available samples per beat can identify. If the PPG diastolic limb tracks that decay even approximately, the per-beat *τ* might index a heartbeat-by-heartbeat refill-like recovery time. We are explicit that this is the weakest of the three physiologically motivated candidate signals: the fingertip diastolic limb is shaped by central compliance, reflection waves, local tone, sensor coupling, and any pre-filter applied by the monitor or by upstream analysis, and is not a capillary-refill measurement.

### 1.6. What we did

For each candidate signal we asked, before computing any downstream association, whether the signal was operating on its intended physiology in the actual data. We drew 10 random instances of the input that produced each per-patient summary value at a fixed seed and inspected whether each instance showed the waveform pattern the signal definition implies. Each instance was reviewed by the human author and, separately, by an on-device multimodal large language model running the same prespecified checklist; disagreements were resolved by the human. A synthetic test against a known time-constant truth checked Signal 3 separately.

## 2. Methods

### 2.1. Data sources and linkage

We used MIMIC-IV version 3.1^10^ as the clinical layer and MIMIC-IV-WDB v0.1.0^9^ as the bedside-monitor waveform layer, both obtained under PhysioNet credentialed access and an executed Data Use Agreement. Downloaded files were SHA-256 verified against the PhysioNet manifest. Each waveform record was linked to MIMIC-IV by overlapping the record’s anchored time window with the patient’s ICU stay. The linkage yielded 147 unique subjects across 148 ICU stays and 149 records. Per-signal admissibility rules reduced the analyzable set to 19 patients for the cuff-anchored signal, 16 patients for the power-ratio signal, and 17 patients for the per-beat exponential signal.

### 2.2. Ethics

MIMIC-IV is distributed by PhysioNet under credentialed access. The Beth Israel Deaconess Medical Center Institutional Review Board approved the creation and sharing of MIMIC-IV with a waiver of informed consent. ^10^ The author completed the required human-subjects training and signed the PhysioNet Data Use Agreement before accessing the data. Because the data are fully de-identified and access is governed by the existing institutional approval and the Data Use Agreement, no additional local Institutional Review Board review was required for this secondary analysis.

### 2.3. Signal preprocessing

The raw PPG was read at the native sampling rate (125 Hz across all records) from the WDB `Pleth` channel. Individual heartbeats were detected with the Elgendi algorithm using the `neurokit2` library. ^25^ The perfusion index (the ratio of pulsatile to non-pulsatile components of the PPG, expressed as a percent) was either taken directly from the monitor where available, or recomputed at one sample per second from the PPG when not: the pulsatile component was the root-mean-square of a band-pass filtered PPG in a 10-second rolling window, the non-pulsatile component was a slow-moving baseline, and PI was their ratio multiplied by 100.

### 2.4. Signal 1: cuff-anchored perfusion-index recovery

#### What we looked for

Around each charted noninvasive blood pressure timestamp the perfusion-index trace should show, if the cuff and the SpO_2_ probe share a limb, a four-phase shape: a stable baseline before the cuff starts; a drop to nearly zero during inflation; a deflation-and-recovery phase as the cuff releases and the small vessels reperfuse; and a return to baseline. The slope and duration of the recovery phase are the candidate microvascular-reactivity readout.

#### Operational definition

A detector ran on a 5-second rolling median of the perfusion-index trace. It required, in this order: a stable baseline window over [`charttime` − 120, `charttime` − 60] seconds; a drop below 50% of baseline that reached a nadir below 20% of baseline; a recovery to at least 85% of baseline sustained for at least 2 seconds; and the nadir falling within an asymmetric window of [− 50, +30] seconds of the charted timestamp (asymmetric because the perfusion drop empirically leads the chart time). The primary classification required the deep-dip phase to last at least 15 seconds; a 10-second threshold was reported as a sensitivity analysis. Routine ICU practice places the cuff and the pulse oximeter probe on opposite limbs, because cuff inflation on the same limb transiently occludes flow to the probe and triggers pulse-wave loss and false low-saturation alarms. ^15,26,27^ The cuff-anchored signal therefore exists, in the recorded data, only on the rare cycles where placement happens to be ipsilateral.

### 2.5. Signal 2: Mayer-band power ratio of the perfusion-index envelope

#### What we looked for

On a stretch of perfusion-index trace several minutes long, the spectrum should contain a clear peak in the slow band (0.05 to 0.15 Hz) corresponding to Mayer-wave oscillations, on a signal that is not drifting or contaminated by motion or by respiration leaking across the band boundary.

#### Why a stationarity check, and what it does

A signal is stationary (in the wide sense relevant here) when its mean, variance, and spectral content are approximately constant across the analysis window. Spectral methods like the Welch periodogram assume this. If the underlying signal is drifting, jumping, or transitioning between physiologic regimes during the window, the resulting spectrum mixes those regimes and the band integrals are not interpretable as the steady-state spectrum of any one regime. The stability check below is a coarse, slope-based attempt to enforce stationarity before estimating the spectrum; the within-window random-instance inspection in Section 2.7 was a second look at whether the assumption held in practice.

#### Operational definition

The 1-Hz perfusion-index trace was segmented into 5-minute windows. A window was admissible if a stability check passed across three 3-minute sub-windows (perfusion-index baseline slope smaller than 3% per hour, heart-rate drift smaller than 5 bpm per hour, respiratory rate within prespecified bounds) and the detected respiratory peak frequency lay outside the low band.

The power spectrum was estimated with the Welch periodogram, a standard signal-processing method that estimates how power is distributed across frequencies by computing the spectra of short overlapping segments and averaging them; we used 256-sample segments with 50% overlap and a Hann taper (a window function that smoothly tapers the segment to zero at its edges to reduce spectral leakage). We chose Welch’s The per-method rather than a single periodogram because biomedical signals are rarely perfectly stationary, and segment-averaging trades a small loss of frequency resolution for a substantial reduction in spectral-estimate variance; this is the convention in the heart-rate-variability and arterial-tone literatures and gives more stable per-window readings. ^20^ Band integrals were 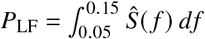 and 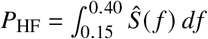, again following standard heart-rate-variability conventions. window signal value was the ratio lf_ratio = *P*_LF_ /(*P*_LF_ +*P*_HF_) . The per-patient summary was the median over admissible windows.

### 2.6. Signal 3: per-beat exponential time constant on the diastolic limb

#### What we looked for

On each PPG beat, the diastolic descent from the dicrotic notch (the small downstroke marking aortic valve closure) to the diastolic minimum should look like a smooth exponential decay, with a recovered time constant in the physiologic range.

#### Operational definition

For every admitted beat the systolic peak, dicrotic notch, and diastolic minimum were localized on the raw PPG. A single-exponential model *y* ( *t*) = A exp (−*t* / *τ*) + *C* was fitted by nonlinear least squares to the raw PPG from the dicrotic notch through the diastolic minimum. The time constant *τ* is the characteristic time over which the value falls by a factor of *e* ≈ 2.72; in a normal arterial system, the diastolic decay time constant is on the order of a few hundred milliseconds. Fit bounds were *τ* ∈ [50, 2000] ms with a goodness-of-fit threshold *R*^2^ ≥ 0.70 required for the beat to be admitted into the per-patient summary. The per-patient summary value was the median *τ* over admitted beats.

#### Pre-fit filtering, and why it matters

An earlier version of the pipeline applied a 0.5–4 Hz Butterworth band-pass to the PPG before the exponential fit. Any linear filter has an intrinsic time constant of its own. A first-order high-pass with corner frequency *f*_*c*_ has *τ*_HP_ = 1 /(2*π f*_*c*_), which for *f*_*c*_ = 0.5 Hz is about 318 ms. An exponential fit applied to a signal that has just passed through such a filter recovers values dominated by *τ*_HP_, not by the underlying physiology. We therefore moved the fit upstream of the band-pass for all results reported below; the synthetic test in Section 3.5 demonstrates the mechanism.

### 2.7. AI-assisted morphology inspection

For each of the three signals we drew 10 random instances of the input that produced the per-patient summary, using `numpy.random.default_rng(GLOBAL_SEED)` with `GLOBAL_SEED = 20260426` fixed in the project source. An instance is a single 5-minute Welch window (Signal 2), a single beat slice from dicrotic notch to diastolic minimum (Signal 3), or a single charted cuff cycle with the surrounding perfusion-index trace (Signal 1). Each instance was rendered as a single plot showing the raw waveform, the model fit or band integrals, and the relevant landmarks, with the per-patient summary value not shown so it could not bias the reader.

A checklist of expected morphology and likely failure modes was written and committed to the project repository before any plot was rendered. For Signal 2 the checklist required: a visible spectral peak above the local noise floor (peak power at least 1.5 times the median power in a reference band at 0.20 to 0.50 Hz) inside the 0.05 to 0.15 Hz Mayer band, on a 5-minute window that was visibly stationary (no drift, dropouts, or motion); the peak not coinciding with a heart-rate subharmonic; the peak not coinciding with the respiratory rate; and the peak not at the two lowest Welch frequency bins (which would indicate residual drift rather than a Mayer-band oscillation). For Signal 3 the checklist required: a visibly mono-exponential decay on the diastolic limb that the model fit tracks; *R*^2^ ≥ 0.80 (tighter than the production threshold of 0.70); *τ* ∈ [50, 1000] ms (tighter than the production upper bound of 2000 ms); and *τ* not within 5% of either fit bound. Failure modes were tallied separately and a failure mode firing on more than 3 of 10 instances was treated as failing the morphology check overall, even if the fixed numerical criteria separately passed.

Each instance was inspected twice. The author inspected and recorded a call on each instance against the checklist. MedGemma 1.5, a multimodal large language model based on Google’s Gemma-3 architecture and fine-tuned for medical content, was then given the same plot and the same checklist by structured prompt and asked to return, in a fixed machine-readable format, a call, a confidence, a short description of what it saw, and a tally of the prespecified failure modes. The model was served locally on the project workstation through an Apple-silicon runtime (oMLX) that exposes an OpenAI-compatible interface; no PhysioNet content was transmitted to any cloud language model service at any point. Decoding was deterministic: the sampling temperature was set to zero, the random seed was fixed, and the output was constrained to the required fields. The model weights were SHA-256 fingerprinted at server start and the prompts were committed to the project repository with cryptographic hashes before any inference. The output constraint took effect only after the model completed an internal reasoning step, and on a subset of windows the model did not complete that step and returned no usable call; these were recorded as non-responses rather than forced into a category. The two sets of calls were compared on the instances the model answered, and the author’s call was used throughout. The language model’s role was an inexpensive second pass against the same prespecified criteria, not an independent scientific judgment. Reporting of this language-model step follows the TRIPOD-LLM guideline. ^28^

### 2.8. Synthetic check for Signal 3

To establish what the per-beat exponential fit recovers when the truth is known by construction, we generated synthetic beats with single-exponential diastolic limbs at three truth values, *τ* ∈ {200, 400, 800} ms, and passed them through two fitting paths: a raw fit on the unfiltered limb, and the same fit applied after the 0.5 Hz high-pass stage used in the earlier pipeline.

### 2.9. Statistical analysis

For Signal 1 the primary estimand was the proportion of evaluable charted cuff cycles that were detector-positive, with subject-clustered nonparametric bootstrap 95% confidence intervals at a fixed random seed; a subject-clustered bootstrap resamples whole patients with replacement (rather than individual cuff cycles) to honor the fact that cycles from the same patient are not independent. A subject-clustered split-half analysis assessed sensitivity of the alignment window. For Signals 2 and 3 we report descriptive distributions and the per-instance morphology counts described above. Cross-signal comparison used Spearman correlation on patients with admitted instances in both signals. Random seeds were fixed for reproducibility: instance sampling used GLOBAL_SEED 20260426, while each bootstrap analysis used its own fixed seed kept separate from the sampling stream (the cross-signal correlation used seed 20260507). No outcome model was fitted; this is a feasibility study.

## 3. Results

### 3.1. Signal 1: the cuff-anchored recovery is rare in MIMIC-IV-WDB

Across 9,224 charted blood pressure cuff cycles in 19 records (19 unique subjects), 8,909 cycles had a usable co-recorded PPG window. A pre-cuff stability check admitted 6,236 cycles in 18 patients. The four-phase occlusion-and-recovery shape was identified on 268 cycles in 15 patients at the primary 15-second recovery threshold, 4.30% of evaluable cycles (Figure 2). A more permissive 10-second threshold yielded 588 cycles in the same 15 patients (9.43% of evaluable). The denominator can also be reported as 268 of 9,224 charted cycles (2.91%), or 15 of 19 patients (79%) with at least one detector-positive cycle. A split-half analysis on a held-out half placed the prespecified alignment window inside both the 95% and 90% data-driven calibration intervals (held-out yield 4.41%, 95% confidence interval 1.52 to 9.56%).

**Figure 2:**
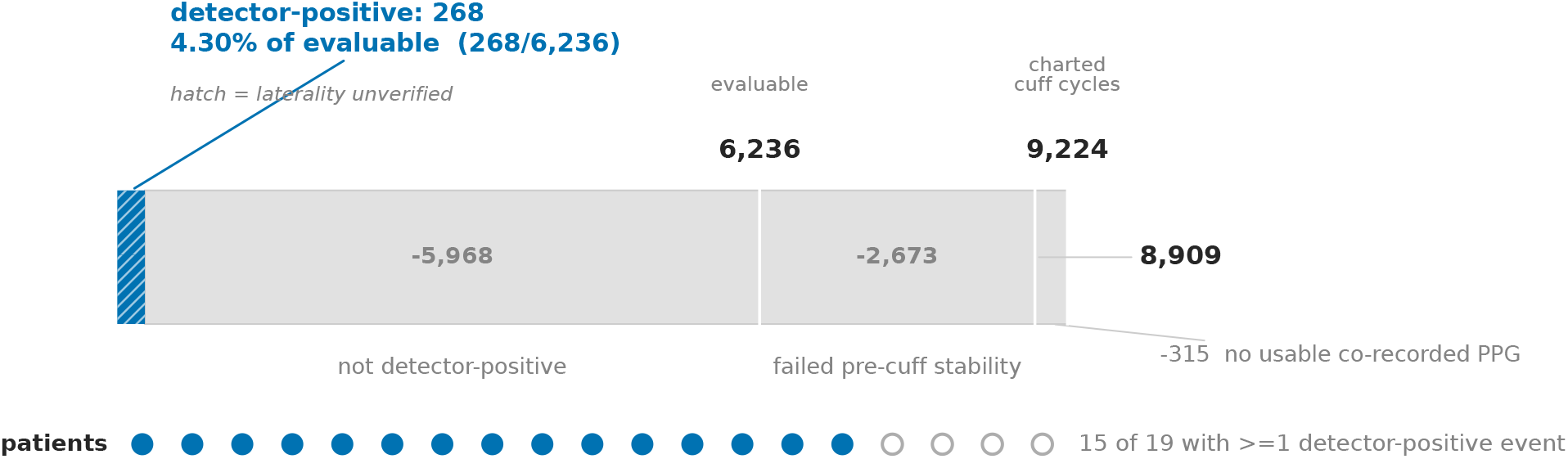
Almost all charted cuff cycles are discarded before a cuff-anchored signal can be read. Of 9,224 charted noninvasive blood pressure cuff cycles, 8,909 had a usable co-recorded photoplethysmogram, 6,236 passed a pre-cuff stability check, and 268 (4.30% of the evaluable cycles, in 15 of 19 patients) showed the four-phase occlusion-and-recovery shape. Bars are drawn to true proportion on a linear scale, so the surviving sliver reflects the actual yield. The diagonal hatch marks that cuff laterality is not recorded in MIMIC-IV-WDB: every detector-positive cycle is a morphology-based estimate, never a confirmed same-limb (ipsilateral) event.

When the morphology was present it was a clean four-phase pattern: a stable pre-cuff baseline, a deep drop in perfusion index, a 15- to 20-second recovery phase, and a return to baseline (Figure 1A). The pattern is consistent with a reactive hyperemia response on the microvascular bed distal to the cuff. The morphology was absent in the great majority of cycles, consistent with the standard-of-care convention of opposite-limb placement. ^15,26,27^

### 3.2. Signal 2: the Mayer-band peak is visible in a minority of random windows

The 1-Hz perfusion-index trace yielded 659 admissible 5-minute Welch windows across 16 patients with at least one admissible window. The per-patient median lf_ratio ranged from 0.280 to 0.711 with cohort median 0.460 and interquartile range 0.417 to 0.532. The number is stable across recomputation at 180, 300, and 600 second window lengths (cohort medians 0.42, 0.46, 0.44 respectively), but the number of patients with any admissible window collapses from 18 to 16 to 12 of 19 as the window grows, which means the apparent stability is conditioned on a shrinking pool.

On visual inspection of 10 random admissible windows the spectral peak the signal is supposed to be reading appeared on a visibly stationary signal in 4 of 10 windows (Figure 3; representative windows in Figure S1, left half), with 1 further window judged borderline. The other 5 windows showed transient perfusion-index dropouts, motion, or baseline drift inside the 5-minute window that the slope-based stability check had not caught, or a peak at the lowest one or two Welch frequency bins (which is what residual drift looks like, not a Mayer oscillation). Respiratory bleed-through across the low-band to high-band boundary appeared in 3 of 10 windows. The local language model returned a usable reading on 6 of the 10 windows and, on the other 4, entered a repetitive internal-reasoning state and returned no call, which we recorded as a non-response. On every window it did read it reported a Mayer peak as present: it agreed with the author on the 4 windows with a clear stationary peak, but it also reported a peak on 2 windows the author had judged artifact, and it never returned a no-peak or indeterminate reading (Figure 4).

**Figure 3:**
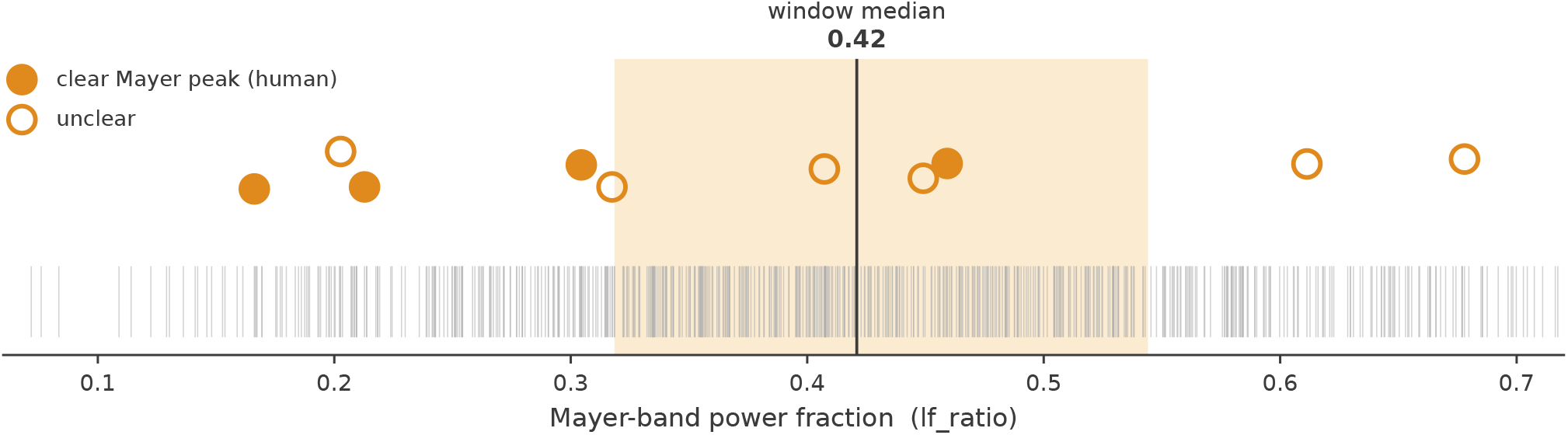
The band-power metric and a reader’s eye disagree about which windows carry a Mayer oscillation. Each tick on the axis is one of 659 admissible 5-minute windows, positioned by its Mayer-band power fraction (lf_ratio, the low-frequency 0.05–0.15 Hz power divided by the low-plus-high-frequency 0.05–0.40 Hz power); the vertical rule is the window-level median (0.42). The ten circles are the windows inspected by eye: filled where the reader judged a clear Mayer peak present, open where unclear. The eye-clear windows sit at or below the median rather than in the high-ratio tail, so a high lf_ratio does not pick out the windows that look like a real oscillation.

**Figure 4:**
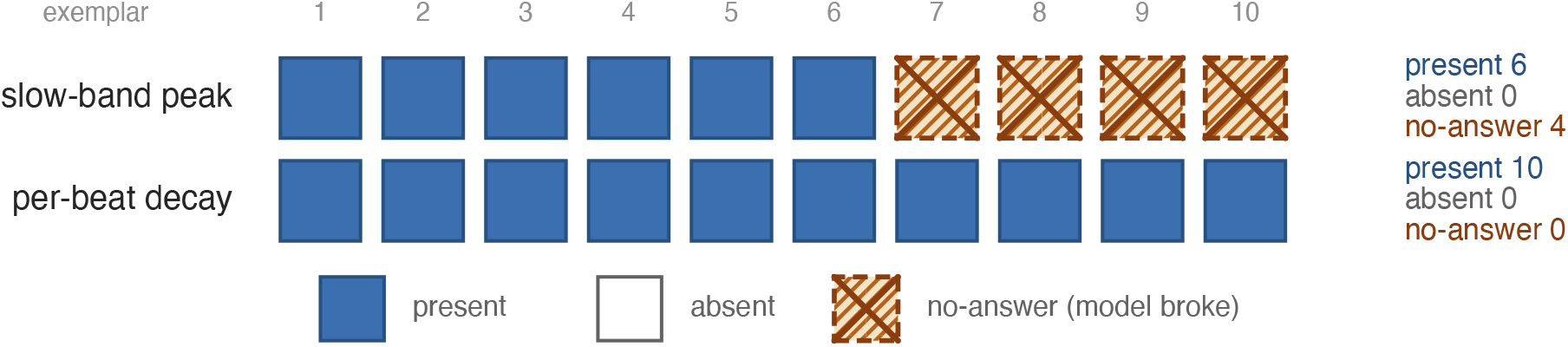
The language model reports the feature present on almost every readable image and breaks on the hardest ones. Each cell is one inspected example: filled where the model called the feature present, struck where it failed to return a usable call (a degenerate non-response), open where it called the feature absent. For the slow-band signal it returned 6 present and 4 non-responses; for the per-beat signal it returned present on all 10 and absent on none. The all-present row is a present-response bias: the model tracked the reader on clear positives but never returned an absent or indeterminate call.

### 3.3. Signal 3: the per-beat fit lands on the heart-rate oscillation in most random beats

The pipeline admitted 237,481 beats across 17 patients. Per-patient median *τ* ranged from 105 to 916 ms with cohort median 280 ms and interquartile range 141 to 518 ms (Figure 5). No patient had a median *τ* stuck at the fit bound. On the 10 random admitted beats, the goodness-of-fit and physiologic-range checks passed in 10 of 10 beats (*R*^2^ ≥ 0.80 and *τ* ∈ [50, 1000] ms in every case). A prespecified cardiac-frequency heuristic, intended to flag fits that may have settled on the heart-rate oscillation rather than on a slow diastolic decay (tell-tale signs are *τ* in the 80–160 ms range and a fit window spanning at most one heart-rate cycle), was met on 7 of 10 beats (Figure S1, right half); at the heart rates common in this cohort a genuinely short physiologic time constant can also fall inside this band, so the heuristic marks beats for scrutiny rather than proving heart-rate entrainment. The local language model returned a usable reading on all 10 beats and reported an exponential decay as present on every one, including the beat the author judged effectively a straight line and the 3 the author judged borderline; setting the borderline beats aside, it agreed with the author on 6 of the 7 remaining beats. As with the slow-band windows, it never returned an absent or indeterminate reading.

**Figure 5:**
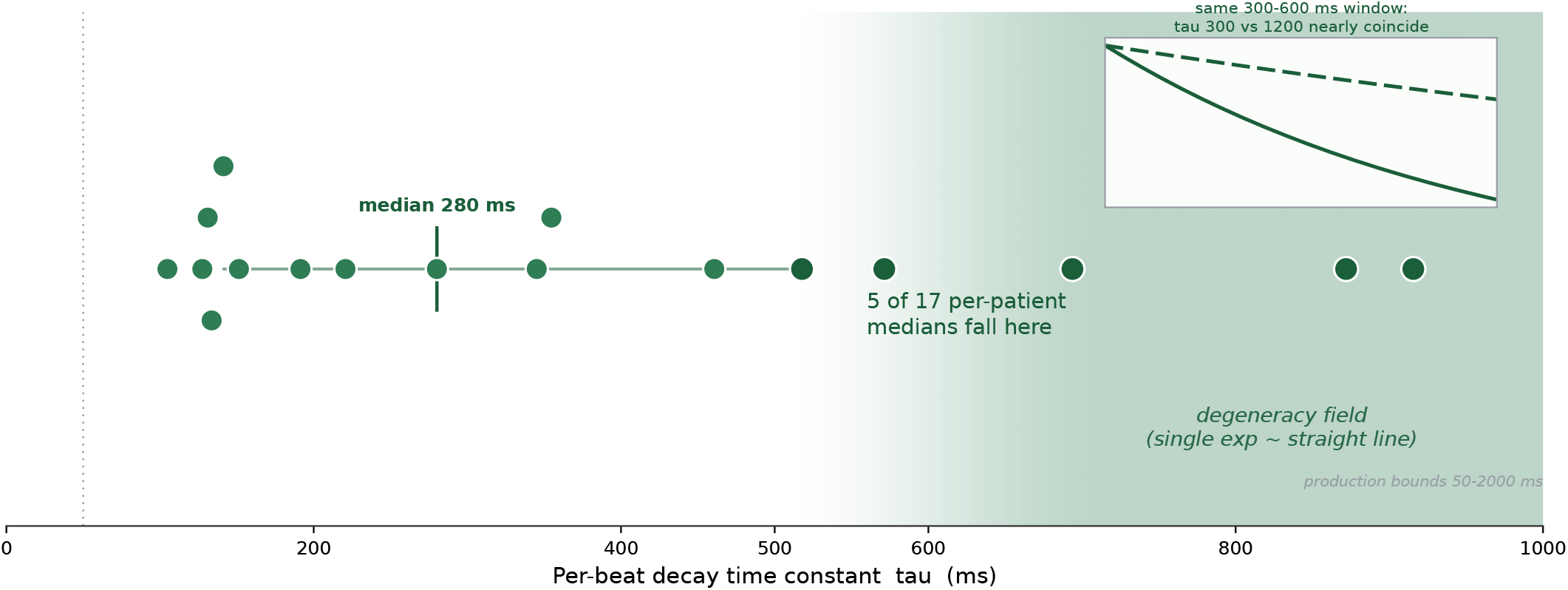
A third of patients fall in a regime where the per-beat fit cannot distinguish an exponential from a straight line. Each dot is one patient’s median per-beat time constant (*τ*, the decay time of the fitted diastolic limb) across 17 patients (cohort median 280 ms, interquartile range 141–518 ms). The shaded field marks where, over a 300–600 ms diastolic window, a single exponential and a straight line through its endpoints become essentially indistinguishable, so a good fit certifies nothing; 5 of 17 patients have a median *τ* inside it. The inset shows two such curves (*τ* of 300 and 1200 ms) nearly coinciding over the fit window.

A separate finding emerged from the per-patient distribution. In 5 of 17 patients (about 29%) the per-patient median *τ* exceeded 500 ms. For *τ* ≥ 500 ms over a 300- to 600-millisecond fit window, the exponential curve and a straight line through its endpoints become essentially indistinguishable; algebraically, the Taylor expansion of A exp(−*t*/*τ*) at small *t*/*τ* is dominated by its linear term. A high *R*^2^ in this regime certifies that some model fits the data, not that the model is specifically an exponential, and the recovered *τ* is then determined by noise rather than by the curvature of the limb. One of the five patients had only 186 admitted beats, the smallest count in the cohort, so the long-*τ* value in that patient is both in the degenerate regime and undersampled.

### 3.4. Cross-signal comparison

If two signals index the same underlying physiology they should correlate across patients. The across-patient Spearman correlation between per-patient median lf_ratio and per-patient median *τ* on the 14 patients present in both tables was *ρ* = −0.38 (95% confidence interval −0.78 to 0.30, 2,000-replicate nonparametric bootstrap, fixed seed 20260507; *p* = 0.17; Figure 6). Within-patient cross-signal correlation on the 8 patients with paired admitted instances had a cohort median of −0.044 (interquartile range −0.288 to 0.233). Both estimates are compatible with no cross-signal association at this cohort size.

**Figure 6:**
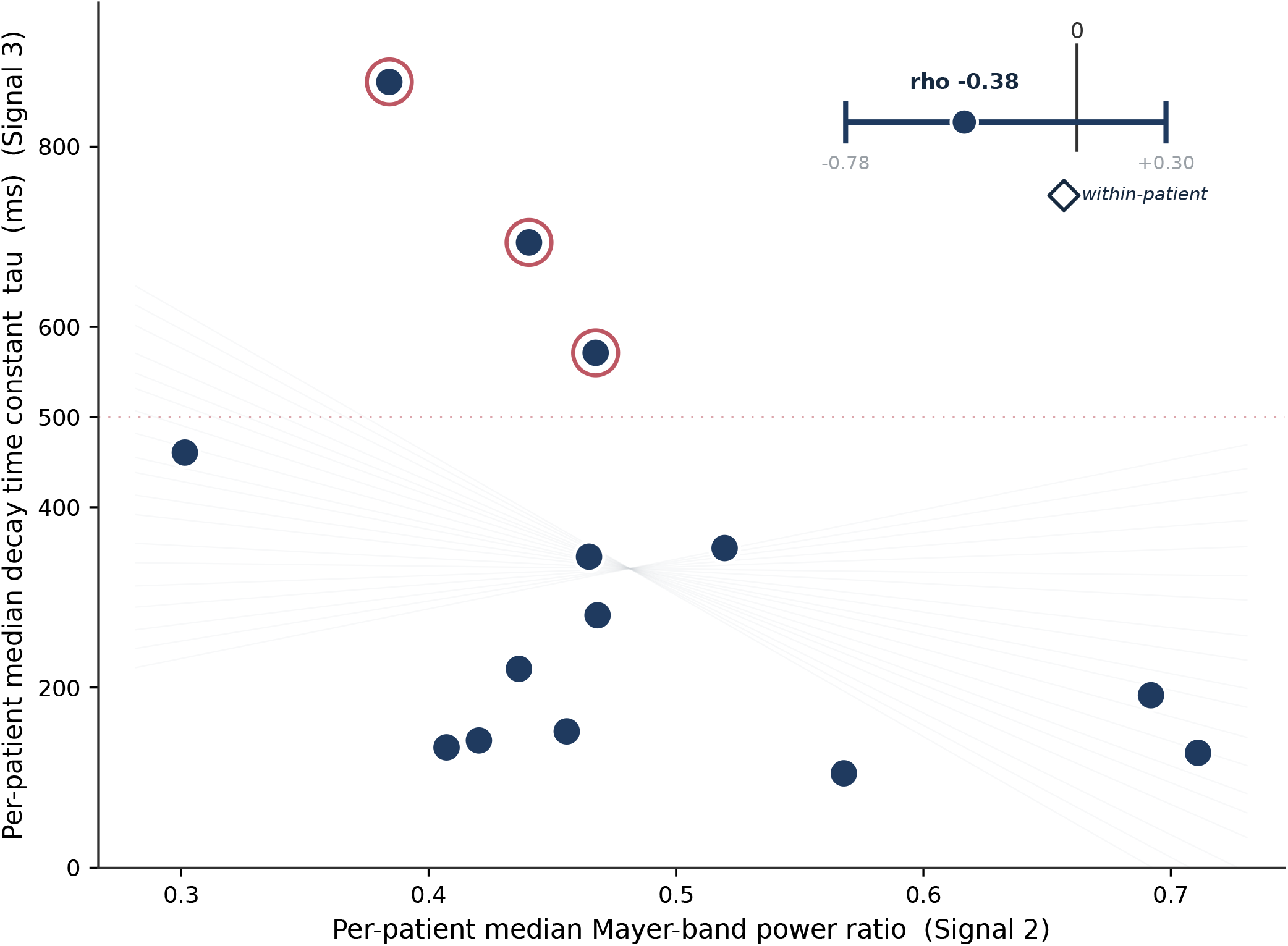
The two slow signals show no reliable across-patient association. Each dot is one of the 14 patients with both a Mayer-band ratio and a per-beat time constant. Rather than a single fitted line, the gray fan shows trends compatible with the data, opening both upward and downward so the uncertainty is visible; the inset marks the Spearman correlation (*ρ* = −0.38, 95% confidence interval −0.78 to 0.30, crossing zero) with the even weaker within-patient value (−0.044). Both are compatible with no association. Red rings flag the patients in the long-time-constant degeneracy regime.

### 3.5. Synthetic check: a high-pass pre-filter implants its own time constant

On synthetic beats with known truth, the raw exponential fit recovered each truth within a 15% tolerance: 200 ms truth →200.88 ms ( +0.44%); 400 ms →413.77 ms ( +3.44%); 800 → ms 904.41 ms ( +13.05%) (Figure 7). The fitting code is not at fault on a clean signal.

**Figure 7:**
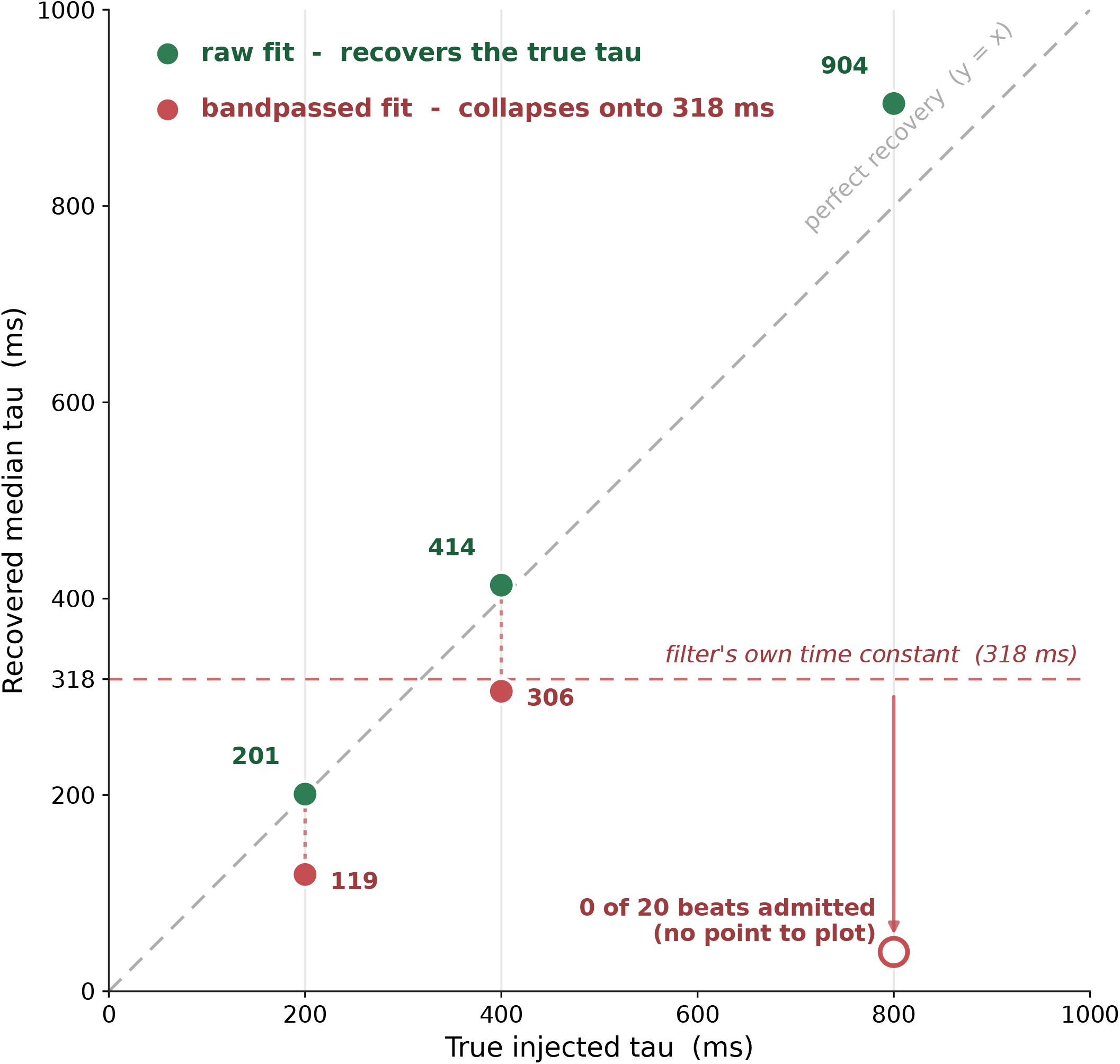
A high-pass pre-filter makes the per-beat fit measure the filter, not the patient. On synthetic beats with a known time constant, the fit applied to the raw signal recovers the truth (200, 400, and 800 ms recovered as 201, 414, and 904 ms, hugging the identity line). The same fit applied after a 0.5 Hz high-pass collapses toward the filter’s own intrinsic time constant of about 318 ms (200 ms read as 119 ms, 400 ms as 306 ms), and at 800 ms no beats survive the filter to fit. Recovering a downstream time constant therefore reflects the pre-processing, not the diastolic decay.

The same fit applied downstream of the 0.5 Hz high-pass did not recover the truth at any setting. At 200 ms truth it returned 118.98 ms ( −40.51%). At 400 ms truth it returned 305.63 ms ( −23.59%). At 800 ms truth the high-pass admitted zero beats and returned no estimate. The 0.5 Hz first-order high-pass has an intrinsic time constant *τ*_HP_ = 1/(2*π*·0.5 ) ≈ 318 ms. The recovered *τ* on filtered input tracks that filter transient rather than the diastolic decay of the underlying beat. The inset of Figure 5 illustrates the long-*τ* identifiability problem directly: a short and a long time constant traced over a typical post-notch window are nearly coincident, so for *τ* ≥ 500 ms the curve and a straight line through its endpoints overlap almost entirely, and a fit can have high *R*^2^ and still not be evidence that the underlying decay is exponential.

## 4. Discussion

We attempted three candidate microvascular-function signals derivable from the photoplethysmogram on MIMIC-IV-WDB v0.1.0 and asked, for each, whether the signal was actually capturing the physiology its definition implied. The cuff-anchored recovery captured the expected shape on 4.30% of evaluable cuff cycles (15 of 19 patients with at least one such cycle), a rarity consistent with the standard-of-care convention that the pulse oximeter probe and the blood pressure cuff are placed on opposite limbs. The Mayer-band power ratio returned a stable cohort-level number but the underlying spectrum was a clear, stationary peak in only 4 of the 10 random windows we inspected. The per-beat exponential fit met its goodness-of-fit threshold in every random beat, but a cardiac-frequency heuristic flagged a possible fit on the heart-rate oscillation, rather than on a slow microvascular decay, in seven of ten random beats, a heuristic that at the heart rates common here also flags some genuinely short physiologic time constants; and in roughly a third of patients the recovered time constant was in a range where the exponential model is mathematically indistinguishable from a straight line. A synthetic test showed that a 0.5 Hz high-pass pre-filter, present in an earlier version of the pipeline, implants its own approximately 318 ms time constant on the recovered value regardless of the underlying truth.

Each of these failure modes is a recognizable family of failure in computational signal work, not a new finding here. A signal can be computed on candidate events whose true class membership the data cannot ground-truth (Signal 1, where the laterality of the cuff and the probe is not recorded in MIMIC-IV-WDB and must be inferred from morphology). A band-power summary can return a stable per-patient number while the underlying spectra are nonstationary, contaminated by motion, or leaked across band boundaries by drift or detrend artifact (Signal 2). A parametric fit can be threshold-stable and return a tightly distributed parameter that reflects fit-window geometry, filter response, or an entrained oscillation rather than the named physiology (Signal 3). A linear filter introduces an intrinsic time constant into every signal it sees, and any downstream parametric fit recovers that time constant when it is comparable to the named target (the synthetic test). The common thread is that each of these failure modes can leave a signal that correlates with severity of illness, because the artifact is correlated with the patient state, even when the signal is not the physiology its definition claims. Cross-feature convergence, robustness across thresholds, and good fit statistics do not separately discriminate signal from artifact.

What we did to detect these failures is simple. We drew 10 random instances of the input that produced each per-patient summary at a fixed seed and looked at the raw signal; the criteria and the failure modes to be tallied were written down before plots were rendered. A second pass on the same plots and the same checklist was run by a multimodal language model served locally on the project workstation. The procedure does not replace synthetic validation, face-validity inspection of curated examples, baseline-comparator analysis, or cross-feature convergence; it sits upstream of all of them. None of those downstream methods can detect that a feature is being computed on artifact if the artifact is itself consistent across patients. The role of the language model here was modest and worth being explicit about. It ran the same prespecified checklist as the human reader on the same anchor-free plots, but its calls showed a marked tendency to report the target pattern as present: on every window and beat for which it returned a usable call, across both signals, it reported the signal as present, and it never returned an absent or indeterminate call. It tracked the human reader on the clearly positive instances but added little for ruling a window or beat out, and on the most ambiguous slow-band windows it returned no call at all. It was an inexpensive second pair of eyes against the same criteria, not an independent rater, and its present-leaning behavior is itself a caution for anyone considering a language model as an automated screen for signal quality. The choice of an on-device model rather than a cloud-hosted service was driven by the PhysioNet Data Use Agreement, which requires that credentialed-access data not be transmitted to third-party services.

The implications for opportunistic computational PPG work are practical. For the cuff-anchored signal, the rate-limiting step is geometry, not signal processing: most cuff cycles in the archive are on the limb opposite the probe, and on those cycles the expected occlusion-reperfusion event does not occur on the recorded signal. A more capable detector would not change that. For the Mayer-band ratio, a slope-based pre-window stability check is not sufficient to ensure within-window stationarity for a Welch-type spectral estimate in critically ill patients; tighter admissibility, paired with random-instance inspection of the windows that pass, would reduce the false-positive rate. For the per-beat exponential signal, the diastolic limb in adult fingertip PPG is too short to identify a slow exponential decay separately from a linear slope in many patients, and any linear pre-filter downstream will dominate the recovered value if its intrinsic time constant is comparable to the named target; a parametric model whose identifiability depends on the relationship between *τ* and the fit window should be evaluated for identifiability before deployment.

The most important limitation of this report is that the random-instance inspection rests on a single human reader paired with a single language model run, on 10 instances per signal. The criteria, the random seed, and the per-instance check tables are committed to the project repository before plots were rendered, so the procedure is reproducible. Inter-rater reliability with a second independent human reader was not estimated and is the obvious next step. The cohort is small: MIMIC-IV-WDB v0.1.0 is an early release that PhysioNet describes as 200 records from 198 patients, and the per-signal analyzable pool was 16 to 19 patients. The cuff-anchored signal was examined on a 19-record pilot inventory and a larger release may yield enough incidentally same-arm cycles for downstream comparison work. The long-*τ* regime in 5 of 17 patients may reflect device, sensor, or filter heterogeneity across the small cohort and was not characterized further here. Within MIMIC-IV-WDB v0.1.0 the laterality of the cuff and the probe is not recorded, so the cuff-anchored signal’s classification is a morphology-based estimate, not a verified placement.

Within MIMIC-IV-WDB v0.1.0, two of the three candidate microvascular signals we attempted did not operate cleanly on their intended physiology in most random instances, and the third was constrained by sensor placement. The downstream associations these signals would support, if computed, would inherit those failures. Drawing 10 random instances of the input that produced each per-patient summary, looking at them against a written-down checklist of expected morphology, and running a language model on the same plots and the same checklist as a second pass, is a small additional cost in computational PPG work and is informative.

## Data Availability

All data referred to in the manuscript are derived from MIMIC-IV (version 3.1) and the MIMIC-IV Waveform Database (MIMIC-IV-WDB v0.1.0), third-party datasets distributed by PhysioNet under credentialed access and a Data Use Agreement. The authors did not generate primary data and do not redistribute the source data, in accordance with the Data Use Agreement; the datasets are available to credentialed researchers at https://physionet.org/content/mimiciv/3.1/ and https://physionet.org/content/mimic4wdb/0.1.0/. The code to reproduce the derived results reported here is available at https://github.com/thomas-landry/ppg-microvascular-feasibility-mimic (archived at https://doi.org/10.5281/zenodo.20534451).

https://physionet.org/content/mimiciv/3.1/

https://physionet.org/content/mimic4wdb/0.1.0/

## AI use disclosure

Two distinct uses of large language models are reported in this work. First, the per-instance morphology second pass described in Methods Section 2.7 used MedGemma 1.5, a multimodal large language model based on Google’s Gemma-3 architecture and fine-tuned for medical content, served locally on the project workstation via an Apple-silicon OpenAI-compatible runtime (oMLX). MedGemma 1.5 read the same anchor-free plots and the same prespecified checklist as the human reader and returned per-criterion calls; the human author’s calls were used throughout, and the instances on which the model returned no usable call were recorded as non-responses. The on-device model received only the rendered morphology plots and the prespecified checklist text; it received no raw waveform samples, no clinical notes, and no record identifiers, and its outputs were limited to the per-criterion calls, confidences, and failure-mode tallies reported here. The model weights were SHA-256 fingerprinted at server start, and the prompts were committed to the project repository with cryptographic hashes before any inference. No PhysioNet content was transmitted to any cloud language model service at any point in this analysis. This local-only design follows the PhysioNet policy on responsible use of credentialed data with online generative-AI services. ^29^ Second, source code drafting, figure scripting, and manuscript drafting were supported by Claude Code (Anthropic); only de-identified derived summaries and code (no raw waveforms or notes) were exchanged. The prespecified morphology checklist, the random seed, and the per-instance check tables were committed to the project repository before any plot was rendered. All scientific judgments, framing, and interpretation are the responsibility of the human author.

## Data and code availability

Source code, the random-sample seed and morphology checklist, the synthetic check, and the test suite are available at https://github.com/thomas-landry/ppg-microva scular-feasibility-mimic (commit `c7b41b9`), archived on Zenodo (https://doi.org/10.5281/zenodo.20534451). The environment lockfile (`pyproject.toml, uv.lock`) and a reproducible figure script are included. MIMIC-IV ^10^ and MIMIC-IV-WDB v0.1.0^9^ are not redistributed; PhysioNet credentialed access and an executed Data Use Agreement are required to reproduce derived feature tables.

## Acknowledgments

MIMIC-IV and MIMIC-IV-WDB are curated by the MIT Laboratory for Computational Physiology and distributed by PhysioNet.

## Supplementary material

**Figure S1:**
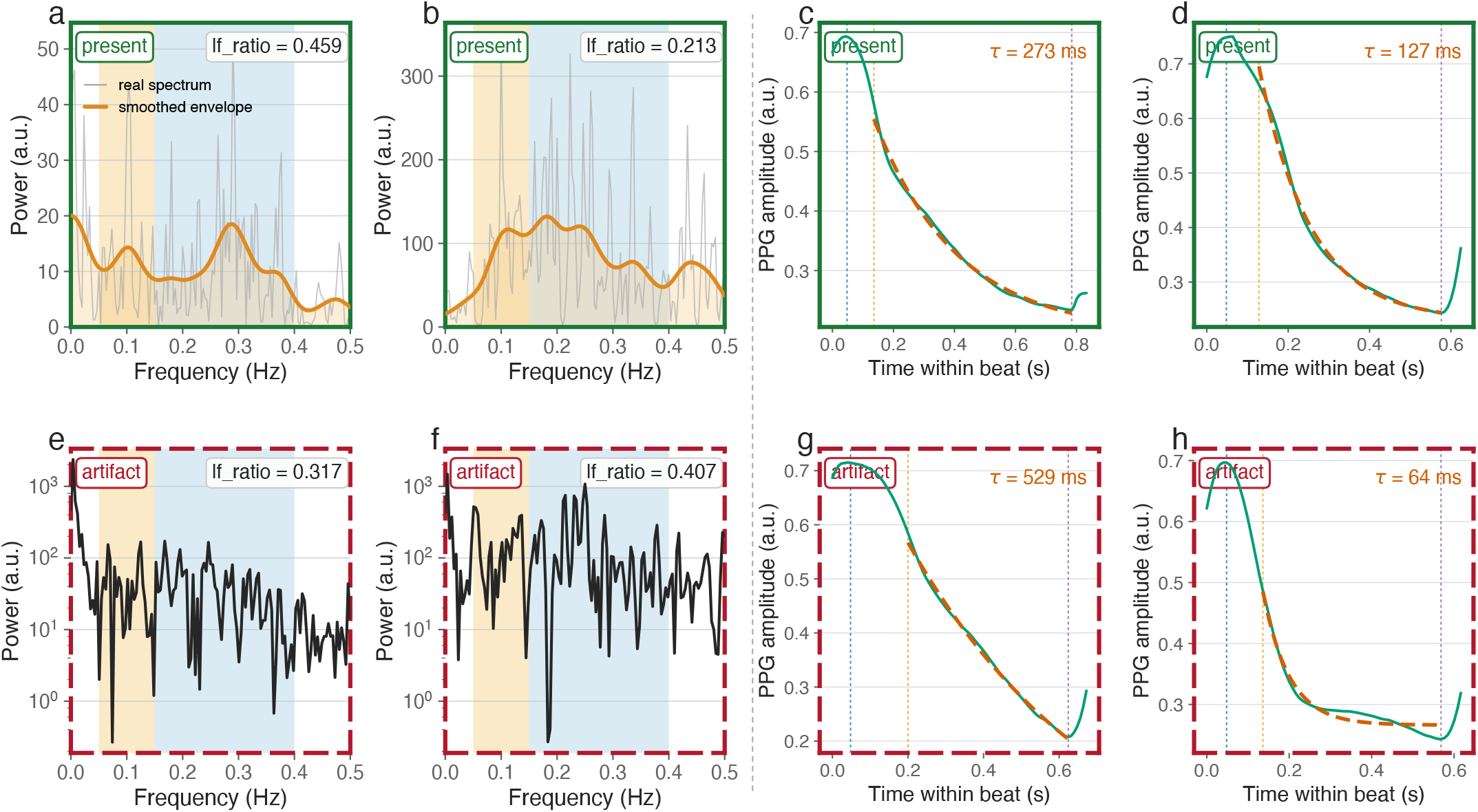
Representative slow-band windows and per-beat decays, present versus artifact. Top row, examples in which the intended feature is visible (solid green borders); bottom row, examples in which the computed value reflects an artifact (dashed red borders). Left half, Signal 2 power spectra with the low- and high-frequency bands shaded and the smoothed envelope overlaid (the labeled value is each window’s lf_ratio); right half, Signal 3 single beats with the fitted exponential and its time constant. All recording-identifying information has been removed; no subject, record, or timing identifiers appear in the figure.

